# Elevated Plasma Complement Factors in *CRB1*-associated Inherited Retinal Dystrophies

**DOI:** 10.1101/2023.11.10.23298334

**Authors:** Lude Moekotte, Joke H. de Boer, Sanne Hiddingh, Aafke de Ligt, Xuan-Thanh-An Nguyen, Carel B. Hoyng, Chris F. Inglehearn, Martin McKibbin, Tina M. Lamey, Jennifer A. Thompson, Fred K. Chen, Terri L. McLaren, Alaa AlTalbishi, Daan M. Panneman, Erica G.M. Boonen, Sandro Banfi, Béatrice Bocquet, Isabelle Meunier, Elfride De Baere, Robert Koenekoop, Monika Ołdak, Carlo Rivolta, Lisa Roberts, Raj Ramesar, Rasa Strupaitė-Šileikienė, Susanne Kohl, G. Jane Farrar, Marion van Vugt, Jessica van Setten, Susanne Roosing, L. Ingeborgh van den Born, Camiel J.F. Boon, Maria M. van Genderen, Jonas J.W. Kuiper

**Affiliations:** Department of Ophthalmology, University Medical Center Utrecht, Utrecht, the Netherlands; Department of Ophthalmology, Leiden University Medical Center, Leiden, the Netherlands; Department of Ophthalmology, Radboud University Medical Center, Nijmegen, the Netherlands; Division of Molecular Medicine, Leeds Institute of Medical Research, University of Leeds, Leeds, United Kingdom; Department of Ophthalmology, St. James’s University Hospital, Leeds, United Kingdom; Centre for Ophthalmology and Visual Science, The University of Western Australia, Perth, Western Australia, Australia; Australian Inherited Retinal Disease Registry & DNA Bank, Department of Medical Technology and Physics, Sir Charles Gairdner Hospital, Nedlands, Western Australia, Australia; St John of Jerusalem Eye Hospital Group, East Jerusalem, Palestine; Department of Human Genetics, Radboud University Medical Center, Nijmegen, the Netherlands; Telethon Institute of Genetics and Medicine, Pozzuoli, Italy; Department of Precision Medicine, University of Campania “Luigi Vanvitelli”, Naples, Italy; Institute for Neurosciences of Montpellier (INM), University of Montpellier, INSERM, Montpellier, France; National Reference Center for Inherited Sensory Diseases, University of Montpellier, CHU, Montpellier, France; Department of Biomolecular Medicine, Ghent University, Ghent, Belgium; Center for Medical Genetics, Ghent University Hospital, Ghent, Belgium; McGill University Health Center (MUHC) Research Institute, Montreal, QC, Canada; Departments of Paediatric Surgery, Human Genetics, and Adult Ophthalmology, McGill University Health Center, Montreal, QC, Canada; Department of Histology and Embryology, Medical University of Warsaw, Warsaw, Poland; Institute of Molecular and Clinical Ophthalmology Basel, Basel, Switzerland; Department of Ophthalmology, University of Basel, Basel, Switzerland; Department of Genetics and Genome Biology, University of Leicester, Leicester, United Kingdom; UCT/MRC Precision and Genomic Medicine Research Unit, Division of Human Genetics, Department of Pathology, Institute of Infectious Disease and Molecular Medicine, Faculty of Health Sciences, University of Cape Town, Cape Town, South Africa; Center of Eye Diseases, Clinic of Ear, Nose, Throat, and Eye Diseases, Institute of Clinical Medicine, Faculty of Medicine, Vilnius University, Vilnius, Lithuania; Institute for Ophthalmic Research, Centre for Ophthalmology, University of Tübingen, Tübingen, Germany; The School of Genetics & Microbiology, The University of Dublin Trinity College, Dublin, Ireland; Department of Cardiology, University Medical Center Utrecht, Utrecht, the Netherlands; Department of Ophthalmology, Het Oogziekenhuis Rotterdam, Rotterdam, the Netherlands; Department of Ophthalmology, Amsterdam UMC, Amsterdam, the Netherlands; Bartiméus, Diagnostic Centre for complex visual disorders, Zeist, the Netherlands

**Keywords:** inherited retinal dystrophy, *CRB1*, proteomics, complement, *CFH*, inflammation

## Abstract

**Objective:** To determine the profile of inflammation-related proteins and complement system factors in serum of *CRB1*-associated inherited retinal dystrophies (*CRB1*-IRDs).

**Design:** A case-control study.

**Subjects, Participants, and/or Controls:** A cohort of 30 Dutch *CRB1*-IRD patients and 29 Dutch healthy controls (HC) (Cohort I), and a second cohort of 123 *CRB1*-IRD patients from 14 countries and 1292 controls (Cohort II) were used in this study.

**Methods:** Quantitative 370-plex targeted proteomics in blood plasma and genotyping of the single nucleotide variant (SNV) rs7535263 in the *CFH* gene.

**Main Outcome Measures:** Plasma concentrations of inflammation-related proteins and the genotype of the SNV rs7535263.

**Results:** *CRB1*-IRD patients showed increased plasma levels of complement system and coagulation cascade proteins compared to healthy controls. Complement Factor I [CFI], Serpin Family D1 [SERPIND1], and Complement Factor H [CFH] were significantly elevated (q<0.05, adjusted for age and sex), which correlated (Pearson’s correlation coefficient >0.6) with higher levels of plasma Complement Component 3 [C3] (q = 0.064). The most enriched pathway in patients was the “Complement cascade” (R-HSA-166658, *Padj* = *P* = 3.03 × 10^-15^). An analysis of the genotype of *CFH* variant rs7535263, which is in close physical proximity to the *CRB1* gene and is associated with other retinal conditions by influencing plasma complement levels, revealed significantly skewed allele distribution specifically in Dutch patients (A allele of rs7535263, odds ratio (OR) [95%CI = 2.85 [1.35-6.02], *P* = 6.19 × 10^-3^), but not in a global case-control cohort (*P* = 0.12). However, *CRB1* missense variants that are common in patients display strong linkage disequilibrium (LD) with rs7535263 in *CFH* in the UK Biobank (D’ = 0.97 for p.(Cys948Tyr); D’ = 1.0 for p.(Arg764Cys)), indicating that genetic linkage may influence plasma complement factor levels in *CRB1*-IRD patients. After accounting for the *CFH* genotype in the proteomic analyses, we also detected significantly elevated plasma levels of Complement Factor H Related 2 [CFHR2] in *CRB1*-IRD patients (q = 0.04).

**Conclusions:** *CRB1*-IRDs are characterized by changes in plasma levels of complement factors and proteins of the innate immune system, which is influenced by common functional variants in the *CFH-CFHR* locus. This indicates that innate immunity is implicated in *CRB1*-IRDs.

## Introduction

The *Crumbs homolog 1 (CRB1)* gene (also known as *RP12*) is a transmembrane protein that functions in the integrity preservation of the photoreceptor layer of the retina. *CRB1* pathogenic variants cause a spectrum of rare autosomal recessive inherited retinal dystrophies (*CRB1*-IRDs) characterized by photoreceptor cell death and early visual impairment.^1–3^ There are many pathogenic variants associated with *CRB1*-IRDs, and the clinical phenotypes vary considerably in severity of disease.^1,3^ Although some correlation between *CRB1* genotype and clinical phenotypes can be found,^4^ overall there is surprisingly little genotype-phenotype correlation, even among cases with identical pathogenic variants.^1,3^ Therefore, it has been hypothesized that phenotypic variation among patients may be caused by factors other than the underlying *CRB1* pathogenic variants.^1^

The presence of vitreous inflammation or cystoid macular edema (CME) in some patients with *CRB1*-IRDs suggests that inflammatory pathways may be involved in the pathophysiology.^5,6^ Indeed, inflammatory genes and complement factors are upregulated in the retinas of rd8 mice homozygous for a *Crb1* gene pathogenic variant.^1,5–9^ Furthermore, we previously showed that patients with *CRB1*-IRDs exhibit changes in their peripheral blood T cells and dendritic cells.^6,10^ These findings suggest that variability of immune responses between patients may contribute to differential clinical phenotypes, but there have been few studies that examine the immune profiles of *CRB1*-IRD patients.^6,10^

A way to study immune profiles is by examining blood proteins, which contribute to immune responses and function by modifying the signaling of cytokines, triggering acute-phase reactions, and activating complement cascades. Targeted proteomics allows the accurate quantification of hundreds of blood proteins in patient samples to determine, in-depth, the immune profile of disease and identify disease-related mechanisms.^11^ This technique has provided insight in the blood proteome of diverse eye conditions such as age-related macular degeneration (AMD), retinopathy of prematurity, uveitis, uveal melanoma, and diabetic retinopathy.^12–18^

In this study, we conducted targeted 370-plex proteomics of blood plasma proteins in *CRB1*-IRD patients and healthy controls to determine the plasma concentrations of inflammation-related proteins. In the same cohort, and in a validation cohort, we determined the *CFH* gene variant rs7535263 to test for associations between protein expression and genotype.

## Methods

### Patients

This study was performed in compliance with the guidelines of the Declaration of Helsinki and has the approval of the local Institutional Review Board (University Medical Center Utrecht (UMCU)). In total, 30 patients were referred to the UMCU from the Amsterdam University Medical Centers, Leiden University Medical Center, Bartiméus Diagnostic Center for complex visual disorders, The Rotterdam Eye Hospital and Rotterdam Ophthalmic Institute, and Groningen University Medical Center for recruitment at the outpatient clinic of the department of Ophthalmology of the UMCU (MEC-14-065). Each patient provided written informed consent before participation and blood collection. The *CRB1*-IRD diagnosis was established by ophthalmic examination, imaging, full field electroretinography, and next-generation sequencing or whole-exome sequencing. Patients were considered to have a molecularly confirmed *CRB1*-IRD in cases where they harbored two or more rare functional variants (i.e., disease-causing *CRB1* variants, such as missense, deletions, loss-of-function, or splice altering variants) affecting both gene copies of *CRB1* (**Supplementary Table 1**).

We excluded patients with systemic inflammatory conditions at the time of sampling and/or patients that received systemic immunomodulatory treatment. At the day of inclusion, patients were examined by visual acuity measurement, and slit lamp examination was performed by an experienced uveitis specialist for assessment of vitreous cells and vitreous haze.

Next, we obtained blood from 29 anonymous age-matched healthy blood bank donors (HC) with no history of ocular inflammatory disease who gave broad written informed consent at the research blood bank of the University Medical Center Utrecht (The “Mini Donor Dienst”).

### Targeted blood proteomics

Venous blood samples were collected in EDTA Lavender Top Tubes (#362084, BD Biosciences, Franklin Lakes, USA) and then centrifuged for 10 min (400g) within an hour of blood collection (**Supplementary Table 2**). Plasma was transferred to a Falcon™ 15 mL Conical Centrifuge Tube and centrifuged for 10 min (1500g), after which the cleared plasma was transferred and stored in Micronic 1.4 mL round bottom tubes (#MP32022, Micronic, Lelystad, the Netherlands) at -80 degrees Celsius. For proteomic analysis of plasma, patient and control samples were randomized over three 96 well plates (#72.1980, Sarstedt, Nümbrecht, Germany) and sealed (#232698, Thermo Fisher, Waltham, USA) prior to shipment to the Olink Proteomic facility at the Erasmus Medical Center, Rotterdam, the Netherlands. We used the *Olink Explore 384 Inflammation II* panel for targeted proteomics of blood plasma. This Olink technology is based on Proximity Extension Assays (PEA) coupled with NGS and is capable of the simultaneous relative quantification of 370 protein analytes.^7^

### TaqMan® SNP genotyping

Genomic DNA of 59 peripheral mononuclear blood samples in RLT Plus lysis buffer (#1053393, Qiagen, Hilden, Germany) was isolated with the AllPrep DNA/RNA/miRNA Universal Kit (#80224, Qiagen, Hilden, Germany). Next, we determined the rs7535263 variant with the TaqMan® SNP genotyping technology (#4351379, Thermo Fisher, Waltham, USA). Genotypes were called using QuantStudio 12K Flex software (Thermo Fisher, Waltham, USA). Linkage disequilibrium (r^2^ or D’) data for rs7535263 in the CEU population of the 1000 genomes were obtained with the *LDproxy tool* with genome build *GRCh37* in *LDlink*.^19^ The estimated recombination rate for CEU (build *GRCh37*) generated by Adam Auton was obtained via the 1000 Genomes ftp site (ftp://ftp.1000genomes.ebi.ac.uk/vol1/ftp/technical/working/20130507_omni_recombination_r <u>ates/</u>).^20^ We calculated the LD between rs7535263 in *CFH* and *CRB1* missense variants p.(Cys948Tyr) and p.(Arg764Cys) in phased whole-genome sequencing data of ∼150,000 individuals of the UK Biobank using the *--ld* function in plink 2.0.^21,22^ The UK Biobank has ethical approval from North West–Haydock Research Ethics Committee (REC reference: 16/NW/0274). Details of the UK Biobank study have been described in detail previously.^23^ This research has been conducted using the UK Biobank Resource under Application Number 24711.

TaqMan® genotyping of rs7535263 was also performed in an independent validation cohort (Cohort II) of 123 *CRB1*-IRD patients from United Kingdom, Australia, Palestine, Italy, France, Belgium, the Netherlands, Canada, Poland, Switzerland, South Africa, Lithuania, Germany, and Ireland (MEC-2010-359) and 1292 Dutch controls from the amyotrophic lateral sclerosis study.^24^ All patients provided written informed consent for genetic testing at the centre of recruitment. *CRB1*-associated genetic diagnoses were made by targeted gene Sanger sequencing or established recently through RP/LCA smMIPs sequencing.^25^ TaqMan® SNP genotyping (#4351379, Thermo Fisher, Waltham, USA) was performed on genomic DNA following the same procedure as described above by the laboratory of the Department of Human Genetics, Radboud University Medical Center, Nijmegen, the Netherlands.

### Statistical analyses

Proteome analyses were performed on *Normalized Protein eXpression* (NPX) units from the output of the Olink analysis in R and R studio (version 4.2.2) using the Olink® Analyze R package.^26,27^ One protein, TNFSF9, had 13 missing data points and 10 proteins did not pass quality control (Olink assay report). These proteins were removed prior to group analyses. The *prcomp* function of R base was used to perform principal component analysis (PCA). Differential expression analysis was performed using a likelihood ratio test (LRT) with age and sex as covariates in the models and we used the *qvalue* package from Bioconductor to correct *P* values for multiple testing.^28^ Over-representation analysis (or enrichment) was performed on proteins with nominal significant differential expression levels (*P*<0.05) with the *ClusterProfiler* R package and using the WikiPathways database as a reference.^29^ The results were plotted using the *dotplot* function of the *enrichplot* R package and the *P*-values from enrichment analysis were adjusted with the Benjamini-Hochberg (BH) procedure, using the *p.adjust* function in R base, and *P*-values (*Padj*) <0.05 were considered statistically significant.^30^ Disease association for rs7535263 was assessed with *Plink* using the Fisher’s exact test and expressed as the odds ratio with a 95% confidence interval. We performed a likelihood ratio test (LRT) to assess the relation between the genotype and the protein expression levels and visualized the results using the *corrplot* R package.^31^ Other figures were generated with the *ggplot2* package.^32^ The explained variance (R2) by the genotype of rs7535263 of plasma proteins was calculated using the squared Pearson correlation coefficient (r^2^) using the *cor* function in R base.

## Results

### Generation of a high-quality plasma proteome of *CRB1*-IRD patients

To explore the composition of inflammatory mediators in the plasma proteome of *CRB1*-IRD patients, we conducted 370-plex targeted proteomics using proximity extension assays combined with *next-generation sequencing* in a cohort of 30 patients and 29 healthy controls. To ensure high-quality data for quantitative proteomic analysis, we first applied a stringent quality control process. We first filtered out low-quality analytes by removing proximity extension assays that did not pass quality control in all plasma samples, resulting in 358 (97%) proteins that passed this filtering step. We detected no outlier plasma samples as shown by the relatively comparable sample median and sample interquartile range (IQR) as well as *Normalized Protein eXpression* (NPX, the protein expression unit which is in Log2 scale) distribution between plasma samples (**Figure 1A and 1B**). However, nine plasma samples exhibited internal controls deviating >0.3 NPX from the plate median which were also removed from the dataset (**Figure 1B**). Principal component analysis on the 50 remaining plasma samples revealed a moderate plate effect that we mitigated by using the *Combat* algorithm to account for batch effects (**Figure 1C and 1D**). As a result, a high-quality proteomic dataset was generated for downstream analysis.

**Figure 1.**
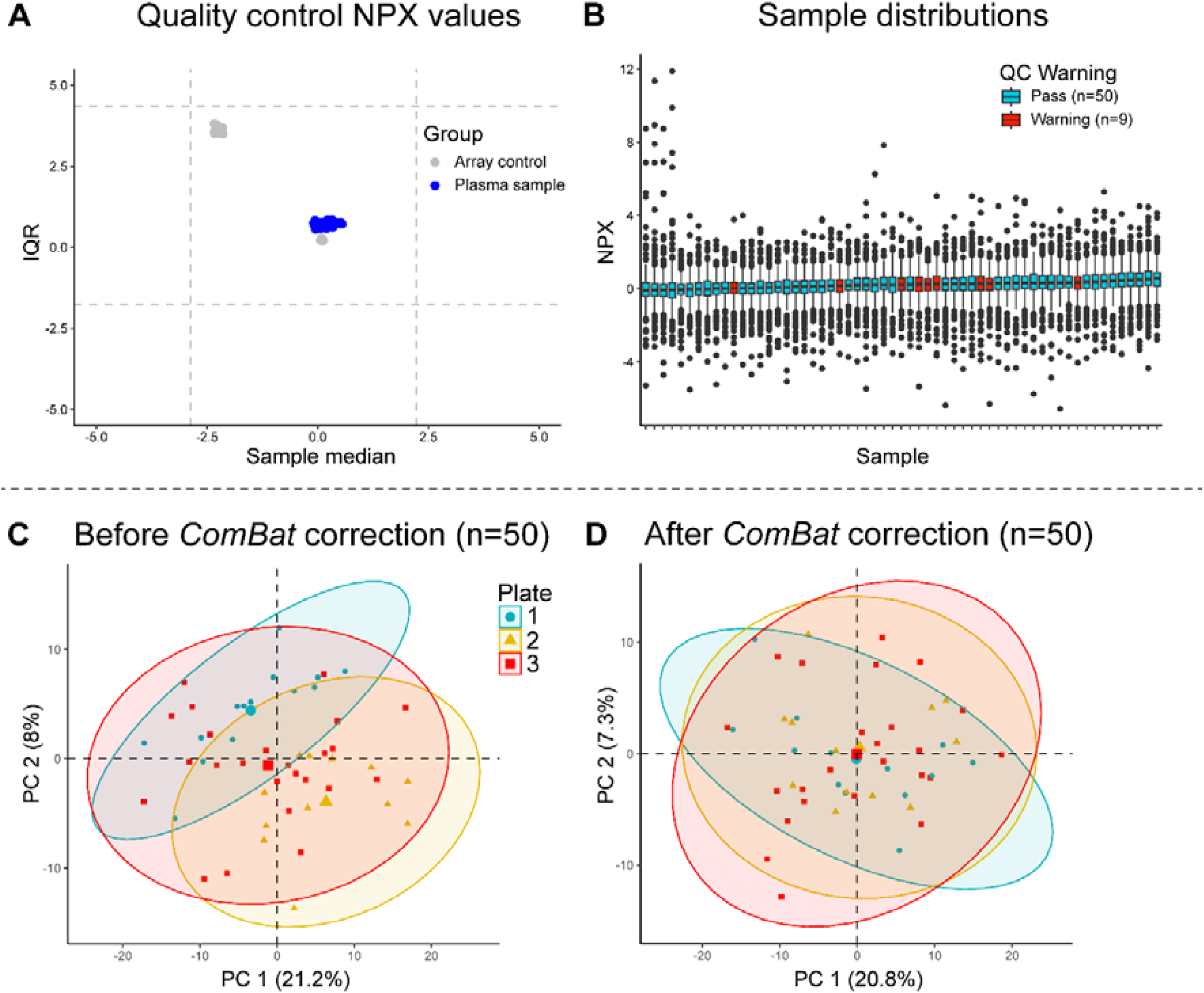
Quality control of plasma proteomic profiling by proximity extension assays combined with *next generation* sequencing. **A)** The median and the interquartile range (IQR) of the distribution of *Normalized Protein eXpression* (NPX) values for each of 59 plasma samples (in blue) measured in this study and internal plate and array controls (in grey). The horizontal and vertical dashed lines indicate +/- 2.5 standard deviations of all sample medians (x-axis) and IQR’s (y-axis). **B**) The sample-wise distributions of NPX values for the Inflammation II panel. Boxplots of NPX values for each plasma sample along the x-axis and NPX values along the y-axis. The center line represents the median. Plasma samples are colored by “QC_Warning” (Warning = >0.3NPX from the median value of all samples on the plate). **C-D**) The first two principal components of 50 plasma samples and 359 proteins before and after batch correction with *Combat*.

### Elevated concentrations of plasma complement factors in *CRB1*-IRD patients

We then compared the plasma proteome of patients to controls using linear models that include age and sex (likelihood ratio test). At a *false discovery rate* of 5% (q<0.05), we detected 10 proteins with significantly different abundance between patients and controls (**Figure 2A** and **Supplementary Table 3**). Among these, elevated levels were observed for Complement Factor I [CFI], Complement factor H [CFH], protein S [PROS1], Serpin family D member 1 [SERPIND1], and A Disintegrin-like And Metalloproteinase with ThromboSpondin type 1 [ADAMTS1] in patients compared to healthy controls (**Figure 2B**). We also detected significantly decreased levels for serine protease 22 [PRSS22] and microfibrillar-associated protein 4 [MFAP4] in *CRB1*-IRD patients.

**Figure 2.**
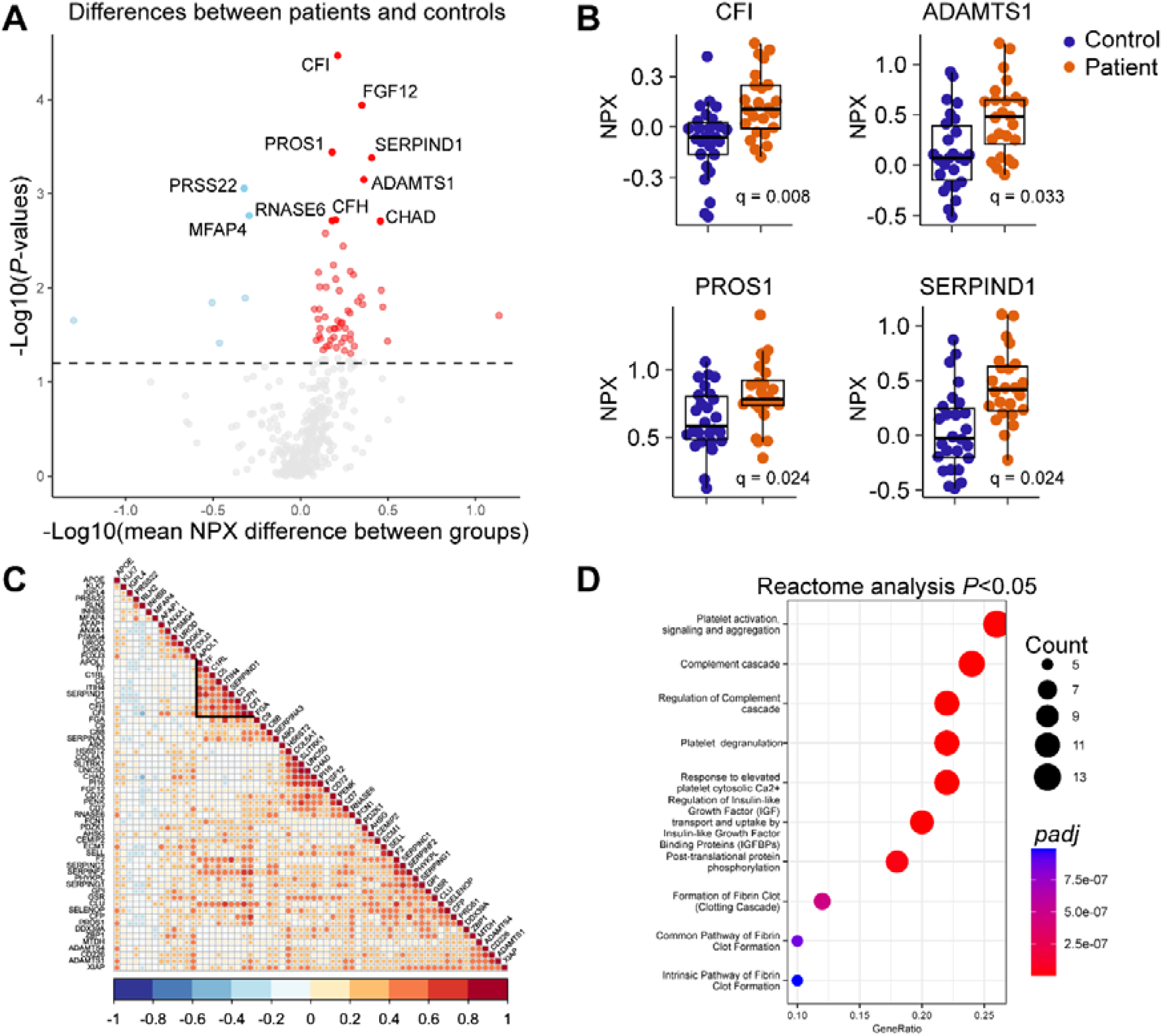
Plasma complement and coagulation proteins are associated with *CRB1*-IRDs. **A)** Volcano plot showing the -Log10(*P*-values) versus the Log2(mean NPX difference between groups) in plasma protein concentrations. Red indicates proteins with a significant increase in protein concentration and blue indicates proteins with a significant decrease in protein concentration in patients compared to controls. The dotted line indicates *P*<0.05. **B)** Scatter plots of the levels of Complement Factor I [CFI], A Disintegrin-like And Metalloproteinase with ThromboSpondin type 1 [ADAMTS1], protein S [PROS1], and Serpin family D member 1 [SERPIND1] in plasma of patients (orange) and controls (blue). The median and the interquartile range (IQR) of the distribution of Normalized Protein eXpression (NPX) values for each plasma sample is shown. **C)** Correlation plot of the 62 proteins most associated with *CRB1*-IRDs (*P*<0.05 from differential expression analysis). Red indicates a positive correlation, white indicates no correlation, and blue indicates a negative correlation between plasma protein analytes. **D)** Dot plot showing the top 10 results from enrichment analysis using the plasma proteins associated with *CRB1*-IRDs as shown in C.

The 62 proteins most associated with *CRB1*-IRD patients (nominal *P*<0.05) showed overall considerable positive correlation in expression levels, with close clustering of CFI, CFH, C3, and SERPIND1 (**Figure 2C**). These 62 proteins were strongly enriched for pathways involving platelet biology, the complement system and coagulation cascades (**Figure 2D** and **Supplementary Table 4**). In detail, the most enriched pathway was the “Complement cascade” (e.g., *Reactome* pathway R-HSA-166658, *Padj* = *P* = 3.03 × 10^-15^, **Figure 2D**). In total, 12/62 *CRB1*-IRD associated proteins were linked to this pathway including the significantly elevated proteins CFH, CFI, and PROS1. In addition, PROS1 and SERPIND1 were also linked to coagulation pathways (i.e., “Formation of Fibrin Clot (Clotting Cascade)”, *Padj* = *P* = 4.69 × 10^-7^). Another enriched pathway included the “Extracellular matrix organization” (R-HSA-1474244, *Padj* = *P* = 5.84 × 10^-3^) based on the significantly altered plasma proteins ADAMTS1 and MFAP4 (**Supplementary Table 4**). Collectively, these results show that elevated levels of complement factors and inflammation-related proteins characterized the plasma proteome of patients with *CRB1*-IRDs.

### The *CFH* genotype is altered in Dutch *CRB1*-IRD patients

On chromosome 1, the *CRB1* gene is located adjacent to the *CFH-CFHR* locus, which is implicated in other retinal conditions by influencing plasma complement protein levels and other proteins detected in our study. Therefore, we hypothesized that linkage between *CRB1* and the *CFH-CFHR* locus could influence the plasma proteome of *CRB1*-IRD patients. To investigate this, we genotyped the common intronic SNP rs7535263 (LD r^2^ = 1.0 with AMD risk variant rs1410996 in CEU population of the 1000 Genomes). The genotype of rs7535263 was shown to have the strongest correlation with the gene expression levels of complement factors H related 1 to 4 (CFHR1-4) in *The Genotype-Tissue Expression* (GTEx) project and is associated with multiple retinal diseases (e.g., AMD, multifocal choroiditis, and serious chorioretinopathy) as well as with other immune mediators on our array, such as CFP.^33–40^

The LD signatures of rs7535263 (r^2^ and D’ with linked SNPs) and genetic recombination rates support that *CRB1* variants and *CFH-CFHR* variants may reside in the same haplotype block (**Figure 3A and 3B**). As expected, the allele frequency of rs7535263 of Dutch controls (A allele of rs7535263 = 0.34) was similar to that of the comparable North-European populations of the 1000 genomes (0.36 in GBR [British in England and Scotland] population). In contrast, the A allele of *CFH*-rs7535263 was significantly higher in frequency in patients compared to controls (A allele of rs7535263, odds ratio (OR) [95%CI = 2.85 [1.35-6.02], *P* = 6.19 × 10^-3^) (**Figure 3C and 3D**). The most common *CRB1* variant in our cohort was the homozygous p.(Met1041Thr) missense variants (*CRB1* c.3122T>C; p.(Met1041Thr)) found in 11/30 patients (37% of patients) which has been previously linked to a large consanguineous pedigree originating from a genetically isolated population in the Netherlands (**Figure 3E and 3F**). While this variant is rare in the general European population (allele frequency = 0.00001 for rs62635656 in gnomAD database), the variant is found relatively more common (>1%) in this specific geographical Dutch area (**Supplementary Table 1**).^41^ We suspected that this deviation in allele frequency may therefore be the result of biological relatedness among cases (i.e., founder effect). All patients in our cohort homozygous for p.(Met1041Thr) were also homozygous for the A allele of rs7535263, which results in an excess of homozygotes for the A allele of the *CFH* variant rs7535263 and provides circumstantial evidence that biological relatedness between cases may likely influence our analysis of plasma complement factors. In a validation cohort of 123 *CRB1*-IRD cases from multiple countries and 1292 Dutch control participants the A allele of rs7535263 was not significantly more frequent in patients compared to controls (A allele of rs7535263, odds ratio (OR) [95%CI = 1.24 [0.95-1.61], *P* = 0.12). However, as in the first cohort, a skewed allele frequency was observed for rs7535263 for the most common *CRB1* variant p.(Cys948Tyr), also a common *CRB1* variant in previous reports,^42^ which suggests linkage between specific variants of *CRB1* and *CFH*. Data from large population databases such as the UK Biobank (UKB) may allow more accurate LD calculation between missense variants in *CRB1* and rs7535263 in the *CFH* gene. To this end, we used phased whole-genome sequencing data from the UK Biobank from ∼150,000 individuals and calculated LD for *CRB1* missense variants. Two *CRB1* missense variants commonly found in *CRB1*-IRD cases with UKB allele frequencies that allow meaningful LD calculations (allele frequency > 0.0001) display strong LD with rs7535263 in *CFH*;^43^ the p.(Cys948Tyr) variant is in LD with the G allele of rs7535263 (D’=0.97, UKB) and the p.(Arg764Cys) T allele is in full LD (D’ =1.0 in UKB) with the A allele of rs7535263. Collectively these results confirm our initial observation that patients with *CRB1* missense variants may co-inherit functional variants in the adjacent *CFH* locus, which should be considered in quantitative analysis of plasma complement factors in *CRB1*-IRD patients.

**Figure 3.**
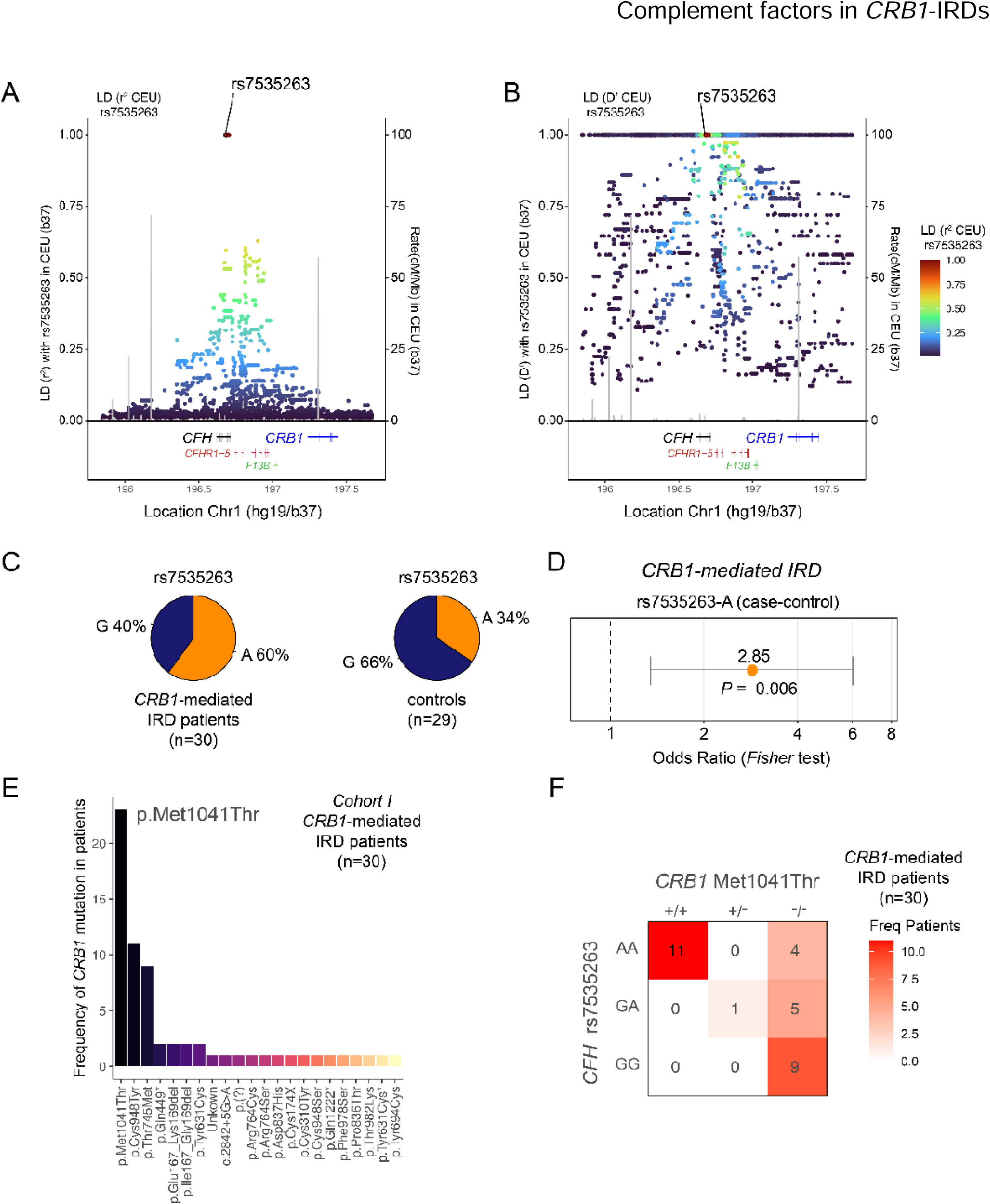
Skewed allelic distribution for a common *CFH* variant in patients with *CRB1*-IRDs. **A-B)** Linkage disequilibrium (r2 metric left, and D’ metric right) for rs7535263 in CEU superpopulation of the 1000 Genomes Project. Genes in the *complement factor H (CFH)* extended locus and *CRB1* are highlighted. **C)** Allele frequency distribution for rs7535263 in cohort 1 in cases and controls. **D)** The odds ratio, 95% confidence interval and *P*-value from Fischer’s exact test for the A allele of rs7535263 in case-control analysis of cohort 1. **E)** The frequency of each *CRB1* pathogenic variant in cohort 1. **F)** Heatmap of the frequency of patients with rs7535263 genotype and *CRB1* Met1041Thr variant dosage in cohort 1.

### *CRB1*-IRD patients show elevated plasma CFHR2 after adjusting for *CFH* genotype

We investigated the effect of rs7535263 on the plasma concentrations of immune mediators in healthy controls. In line with previous studies, the genotype of rs7535263 explained a significant proportion of the plasma levels of several proteins encoded by genes in the extended *CFH-CFHR* locus, including Complement factor H related protein 2 (CFHR2) (R2 = 0.36), and CFHR4 (R2 = 0.24), but not CFH (R2 = 0.003) (**Supplementary Table 5 with R2 values**), indicating that the genotype of this SNP may interfere with our analysis of some proteins in *CRB1* patients. We therefore repeated our proteomic analysis for cohort 1 while correcting for the genotype of rs7535263 (along with sex and age). This analysis revealed that the association with the 10 significantly altered plasma proteins (q<0.05) in our initial group comparison were not significantly influenced by the genotype, such as CFI (q_rs7535263_ = 0.003), and CFH (q_rs7535263_ = 0.06) (**Figure 4A and 4B, Supplementary Table 3**) and all 10 remained within an FDR of 7% (q<0.07). However, adjusting for rs7535263 revealed additional proteins associated with *CRB1*-IRDs, most likely by improving statistical power by reducing residual variance. Among these plasma proteins was CFHR2 (q_rs7535263_ = 0.04), further supporting dysregulation of complement pathways in *CRB1*-IRDs (**Figure 4A and 4B**). Correcting for rs7535263 also revealed significant association (q_rs7535263_<0.05) with the DNA binding protein ZBP1, and RNA helicase DDX39A, which was previously also associated with the genotype of rs7535263 in multifocal choroiditis patients.^38^

**Figure 4.**
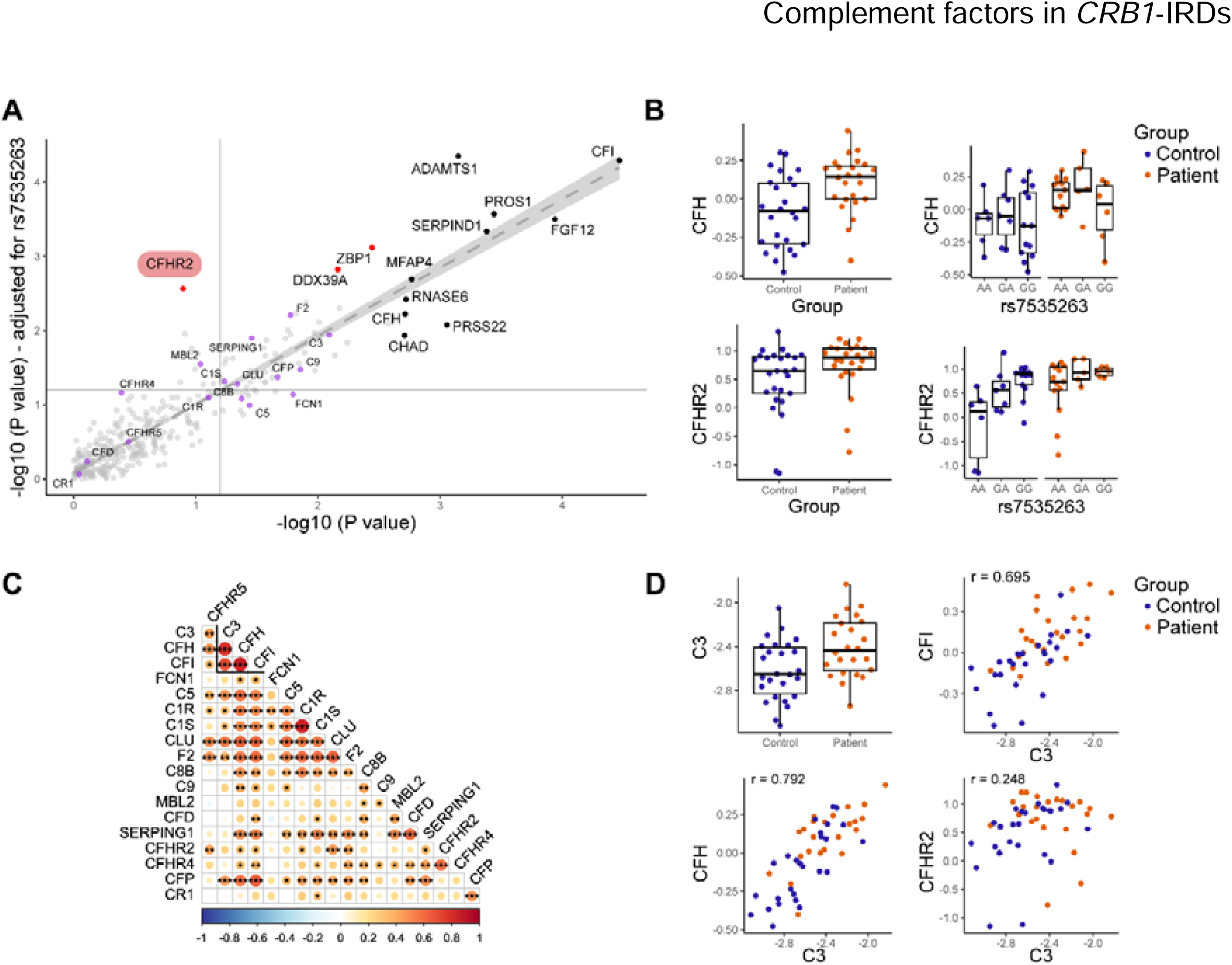
Elevated CFI and CFH levels correlate with plasma C3 in *CRB1*-IRDs. **A)** Results from differential expression analysis before (x-axis) and after (y-axis) corrected for the genotype of rs7535263. The -Log10(*P*-value) for each protein analyte is shown. Red indicates proteins that became statistically significantly different between cases and controls after adjusting for the genotype of rs7535263 and the color black indicates the 10 significant plasma proteins (q<0.05) in our initial group comparison. Purple highlights plasma proteins involved in complement pathways. The dotted line indicates *P*<0.05. **B)** Boxplots showing the levels for complement factor H [CFH] and complement factor H related 2 [CFHR2] in cases and controls in bulk and stratified by the genotype of *CFH* variant rs7535263. **C)** Correlation plot of the 18 proteins involved in complement pathways. Red indicates a positive correlation, white indicates no correlation, and blue indicates a negative correlation between plasma protein analytes. **D)** Boxplot and correlation plots of the levels of Complement Factor I [CFI], CFH, CFHR2, and complement component 3 [C3] in plasma in patients (orange) and controls (blue).

The identified complement factors CFI, CFH, and CFHR2 all target C3,^44,45^ an important downstream effector of the complement system which was also increased in *CRB1*-IRD patients, albeit at nominal significance (q = 0.064) (**Figure 4D**). Based on the profile of complement factors in our array (n = 18), we performed a correlation analysis to identify the factors associated with plasma C3 levels in our cohort. This analysis revealed that CFH and CFI, but not CFHR2, strongly correlated with the levels of plasma C3 (**Figure 4C and 4D**).

## Discussion

In this study, we examined the plasma proteome of patients with *CRB1*-IRDs, whose levels of inflammation-related proteins and complement factors were notably elevated compared to healthy controls. A remarkable observation was the detection of elevated CFH and CFHR2 proteins in *CRB1*-IRDs because elevated plasma levels of CFHR2 have been linked with other eye conditions, including AMD and multifocal choroiditis patients.^12,33,34,38,46^ CFH/CFHR proteins are all involved in the regulation of the complement component C3,^45,47^ which was supported by our observation of strong correlation between CFI and CFH with C3 in plasma. Our results are also in line with animal models of IRDs (rd10 mice), which show an increase in C3 protein levels and C3 activation, as well as an increase of mRNA of *cfh*, and *cfi* in the retina.^48^ While C3 is important for complement activation and can cause microglial activation and photoreceptor cell injury,^49^ it has also been shown to mediate retina-protective signaling in other murine models of retinal degeneration but will require human trials with (intravitreal) anti-C3 therapy to elucidate its potential as a therapeutic target for *CRB1*-IRDs.^48,50,51^ Regardless, the increased levels of complement factors and inflammation-related proteins that we found in patients support immune-mediated pathways involved in *CRB1*-IRDs.

Among the plasma proteins that were decreased in plasma of *CRB1*-IRD patients was the extracellular protein MFAP4 that is ubiquitously expressed and plays a role in intracellular interactions and cell adhesion. Macrophages drive local immune responses in retinal degeneration^52,53^ and MFAP4 loss affects macrophage differentiation in animal models.^54^ The elevated plasma levels of ADAMTS1 detected in *CRB1*-IRD patients may be related to angiogenic responses, due to the vascular impairment secondary to degenerative changes in the retina.^55–58^ The ADAMTS1 enzyme is an angiogenic matrix-modifying enzyme that is upregulated by pro-inflammatory conditions in the retinal pigment epithelium (RPE).^59^ We also detected elevated levels of PROS1, a protein involved in the homeostasis of RPE. Whole-exome sequencing has previously identified homozygous pathogenic variants in the *PROS1* gene as causing retinal dystrophy in two unrelated families,^60^ potentially disrupting the vitamin K-dependent activities of PROS1, leading to RPE degradation. Further investigation is needed to determine whether the elevated levels of PROS1 are caused by retinal damage or by other biological events involving PROS1.

While the plasma proteome may reflect inflammation at the retinal level, it may also be influenced by functional variants in complement factor coding genes. The *CRB1* gene is located on chromosome 1 immediately downstream of the *CFH* locus. SNPs in the *CFH* locus have been associated with several retinal conditions in which CFH/CFHR proteins were also altered, including AMD, central serous chorioretinopathy, and multifocal choroiditis,^33–39^ indicating that this locus could be an important disease-modifying locus for eye conditions.

Using TaqMan® SNP genotyping technology, we found evidence for skewed distribution of functional variants in the *CFH* gene which may predispose to altered complement factor levels in *CRB1*-IRDs or obscure relevant disease associations with plasma proteins, as demonstrated by our work. The significant skewing of the common *CFH* variants across *CRB1* pathogenic variants in our Dutch cohort is likely the result of frequent consanguineous marriages in geographically more isolated populations, such as the strict geographical location of the p.(Met1041Thr) in the Netherlands and predisposes to homozygosity of harmful recessive alleles. Genotype frequencies, Hardy-Weinberg equilibrium assumptions, and allele frequencies can be affected by relatedness and our study demonstrates that addressing relatedness is crucial to ensure the accuracy of genotype-disease relationships and assessment of quantification of the complement factors in *CRB1*-IRDs and other family-based monogenic conditions. We believe this genetic “confounding” of common variants is currently strongly underappreciated in molecular profiling studies of *CRB1*-IRDs.

While we show that in a second global cohort there is no association with the *CFH/CFHR* locus in *CRB1* cases in general, data from the well-powered UK Biobank demonstrated linkage between variants in *CFH* and *CRB1*. Several key missense variants (e.g., p.(Cys948Tyr)) are nearly exclusively inherited with one of the two alleles of rs7535263, which tag functionally distinct *CFH/CFHR* haplotypes and predispose to altered composition of plasma complement factors.^11,12^ This linkage is expected from their proximity on the genome and LD signatures in the general population and has important implications for future research and treatment modalities: 1) Our study demonstrates that complement factor components are involved in *CRB1*-IRDs, but that relative biological relatedness (i.e., founder effects) between individuals or linkage with functional variants in the *CFH* locus should be considered in follow-up studies of complement dysregulation in human studies of *CRB1*-IRDs. 2) While we demonstrate that correcting for the genotype of rs7535263 revealed an otherwise obscured increase in CFHR2 levels in patients, it is not unlikely that additional CFH(R) gene variants that genetically predispose to altered complement factor levels contribute to the pathophysiology of *CRB1*-IRDs. The limited sample size of our study and the lack of probing for other CFH-related factors encoded by genes upstream of the *CRB1* locus (e.g., CFHR1 and CFHR3 levels correlate with genotype of rs7535263 according to *gtexportal*) may have resulted in an underestimation of the differences in the levels of complement factors in *CRB1*-IRD patients compared to controls. 3) Many more common variants in immune genes are likely involved in disease-modifying pathways that may influence the severity and disease course of *CRB1*-IRDs. The use of genome-wide association studies using well defined clinical endpoints, preferably based on microperimetry,^61^ may aid in the discovery of these common variants and could improve genotype-phenotype correlations to better predict disease course, and perhaps lead to therapeutic modalities that can alter the disease course. The latter may be also of interest for emerging *CRB1* gene-augmentation therapy or mutational correction by allelic substitution via CRISPR/Cas9-mediated homology-directed repair or base editing,^62,63^ because these therapeutic approaches are accompanied by immune activation when administered intravitreally or subretinally.^64^ It would be important to determine to what extent the inflammatory markers identified in this study may help understand the side effects of gene therapy, such as increased chorioretinal atrophy that has recently been described after gene therapy using *voretigene neparvovec* for *RPE65*-associated retinal dystrophies.^65,66^ A major source of plasma complement components is the liver, but complement proteins are also produced and expressed in the retina, as well as by immune cells and various tissues. Complement activation may occur in the retina due to loss of tissue integrity caused by *CRB1* dysfunction, and modulating the complement system without addressing this primary cause may not have a clinically significant impact on the progression of the disease. Regardless, a complement-targeted therapy may be useful as a combination therapy to minimize adverse reactions from *CRB1* gene correcting strategies in retinal degeneration, and potentially even in other gene therapy strategies for IRDs. In future studies, it might be informative to determine the immune profile of patients prior to commencing gene therapy interventions, to determine to what extent complement activation and plasma concentrations influence outcome in patients.

In conclusion, our results implicate the involvement of the complement cascade in *CRB1*-IRDs. For future research, gene association studies of all SNPs in the genome with a categorization of patients based on disease severity may provide more insight into the pathophysiology of *CRB1*-IRDs, and IRDs in general.

## Data Availability

All data produced in the present study are available upon reasonable request to the authors

## Acknowledgements

Dr. N.H. ten Dam-van Loon, Dr. J. Ossewaarde-van Norel, and Dr. V. Koopman-Kalinina Ayuso for their assistance performing slit lamp examination and assessment of vitreous cells and vitreous haze on all included patients.

## References

1. Bujakowska K, Audo I, Mohand-Säid S, et al. CRB1 mutations in inherited retinal dystrophies. Hum Mutat 2012;33:306. Available at: /pmc/articles/PMC3293109/ [Accessed August 7, 2022].

2. Den Hollander AI, Heckenlively JR, van den Born LI, et al. Leber congenital amaurosis and retinitis pigmentosa with Coats-like exudative vasculopathy are associated with mutations in the crumbs homologue 1 (CRB1) gene. Am J Hum Genet 2001;69:198–203. Available at: https://pubmed.ncbi.nlm.nih.gov/11389483/ [Accessed September 6, 2023].

3. Den Hollander AI, Davis J, Van Der Velde-Visser SD, et al. CRB1 mutation spectrum in inherited retinal dystrophies. Hum Mutat 2004;24:355–369. Available at: https://pubmed.ncbi.nlm.nih.gov/15459956/ [Accessed September 6, 2023].

4. Talib M, Van Cauwenbergh C, De Zaeytijd J, et al. CRB1-associated retinal dystrophies in a Belgian cohort: genetic characteristics and long-term clinical follow-up. British Journal of Ophthalmology 2022;106:696–704. Available at: https://bjo-bmj-com.proxy.library.uu.nl/content/106/5/696 [Accessed October 3, 2023].

5. Hettinga YM, van Genderen MM, Wieringa W, et al. Retinal Dystrophy in 6 Young Patients Who Presented with Intermediate Uveitis. Ophthalmology 2016;123:2043–2046. Available at: https://pubmed.ncbi.nlm.nih.gov/27157150/ [Accessed February 21, 2023].

6. Verhagen F, Kuiper J, Nierkens S, et al. Systemic inflammatory immune signatures in a patient with CRB1 linked retinal dystrophy. Expert Rev Clin Immunol 2016;12:1359–1362. Available at: https://pubmed.ncbi.nlm.nih.gov/27690673/ [Accessed February 21, 2023].

7. Watson CM, El-Asrag M, Parry DA, et al. Mutation screening of retinal dystrophy patients by targeted capture from tagged pooled DNAs and next generation sequencing. PLoS One 2014;9. Available at: https://pubmed.ncbi.nlm.nih.gov/25133751/ [Accessed August 8, 2022].

8. De Castro-Miró M, Pomares E, Lorés-Motta L, et al. Combined genetic and high-throughput strategies for molecular diagnosis of inherited retinal dystrophies. PLoS One 2014;9. Available at: https://pubmed.ncbi.nlm.nih.gov/24516651/ [Accessed August 8, 2022].

9. Alves CH, Wijnholds J. Microglial Cell Dysfunction in CRB1-Associated Retinopathies. Adv Exp Med Biol 2019;1185:159–163. Available at: https://link-springer-com.proxy.library.uu.nl/chapter/10.1007/978-3-030-27378-1_26 [Accessed August 8, 2022].

10. Moekotte L, Kuiper JJW, Hiddingh S, et al. CRB1-Associated Retinal Dystrophy Patients Have Expanded Lewis Glycoantigen-Positive T Cells. Invest Ophthalmol Vis Sci 2023;64:6. Available at: https://pubmed.ncbi.nlm.nih.gov/37792335/ [Accessed October 10, 2023].

11. Sun BB, Maranville JC, Peters JE, et al. Genomic atlas of the human plasma proteome. Nature 2018;558:73–79. Available at: https://pubmed.ncbi.nlm.nih.gov/29875488/ [Accessed September 7, 2023].

12. Emilsson V, Gudmundsson EF, Jonmundsson T, et al. A proteogenomic signature of age-related macular degeneration in blood. Nat Commun 2022;13. Available at: https://pubmed.ncbi.nlm.nih.gov/35697682/ [Accessed September 7, 2023].

13. Lynch AM, Wagner BD, Mandava N, et al. The Relationship of Novel Plasma Proteins in the Early Neonatal Period With Retinopathy of Prematurity. Invest Ophthalmol Vis Sci 2016;57:5076–5082. Available at: https://pubmed.ncbi.nlm.nih.gov/27679852/ [Accessed September 7, 2023].

14. Kuiper JJW, Verhagen FH, Hiddingh S, et al. A Network of Serum Proteins Predict the Need for Systemic Immunomodulatory Therapy at Diagnosis in Noninfectious Uveitis. Ophthalmology science 2022;2. Available at: https://pubmed.ncbi.nlm.nih.gov/36245752/ [Accessed September 7, 2023].

15. Achten R, van Luijk C, Thijs J, et al. Non-Infectious Uveitis Secondary to Dupilumab Treatment in Atopic Dermatitis Patients Shows a Pro-Inflammatory Molecular Profile. Ocul Immunol Inflamm 2023. Available at: https://pubmed.ncbi.nlm.nih.gov/36854134/ [Accessed September 7, 2023].

16. Wennink RAW, Ayuso VK, Tao W, et al. A Blood Protein Signature Stratifies Clinical Response to csDMARD Therapy in Pediatric Uveitis. Transl Vis Sci Technol 2022;11. Available at: https://pubmed.ncbi.nlm.nih.gov/35103800/ [Accessed September 7, 2023].

17. Wierenga APA, Cao J, Mouthaan H, et al. Aqueous Humor Biomarkers Identify Three Prognostic Groups in Uveal Melanoma. Invest Ophthalmol Vis Sci 2019;60:4740–4747. Available at: https://pubmed.ncbi.nlm.nih.gov/31731294/ [Accessed September 7, 2023].

18. Haq Z, Yang D, Psaras C, Stewart JM. Sex-Based Analysis of Potential Inflammation-Related Protein Biomarkers in the Aqueous Humor of Patients With Diabetes Mellitus. Transl Vis Sci Technol 2021;10:1–8. Available at: https://pubmed.ncbi.nlm.nih.gov/34003946/ [Accessed September 7, 2023].

19. Machiela MJ, Chanock SJ. LDassoc: an online tool for interactively exploring genome-wide association study results and prioritizing variants for functional investigation. Bioinformatics 2018;34:887–889. Available at: https://pubmed.ncbi.nlm.nih.gov/28968746/ [Accessed September 7, 2023].

20. Anon. Index of /vol1/ftp/technical/working/20130507_omni_recombination_rates. Available at: https://ftp.1000genomes.ebi.ac.uk/vol1/ftp/technical/working/20130507_omni_recombination_rates/ [Accessed October 10, 2023].

21. Chang CC, Chow CC, Tellier LCAM, et al. Second-generation PLINK: rising to the challenge of larger and richer datasets. Gigascience 2015;4. Available at: https://pubmed.ncbi.nlm.nih.gov/25722852/ [Accessed October 4, 2023].

22. Hofmeister RJ, Ribeiro DM, Rubinacci S, Delaneau O. Accurate rare variant phasing of whole-genome and whole-exome sequencing data in the UK Biobank. Nat Genet 2023;55:1243–1249. Available at: https://pubmed.ncbi.nlm.nih.gov/37386248/ [Accessed October 10, 2023].

23. Yeung MW, Wang S, Van De Vegte YJ, et al. Twenty-Five Novel Loci for Carotid Intima-Media Thickness: A Genome-Wide Association Study in >45D000 Individuals and Meta-Analysis of >100D000 Individuals. Arterioscler Thromb Vasc Biol 2022;42:484–501. Available at: https://www-ahajournals-org.proxy.library.uu.nl/doi/abs/10.1161/ATVBAHA.121.317007 [Accessed October 10, 2023].

24. van Rheenen W, van der Spek RAA, Bakker MK, et al. Common and rare variant association analyses in amyotrophic lateral sclerosis identify 15 risk loci with distinct genetic architectures and neuron-specific biology. Nat Genet 2021;53:1636–1648. Available at: https://pubmed.ncbi.nlm.nih.gov/34873335/ [Accessed November 2, 2023].

25. Panneman DM, Hitti-Malin RJ, Holtes LK, et al. Cost-effective sequence analysis of 113 genes in 1,192 probands with retinitis pigmentosa and Leber congenital amaurosis. Front Cell Dev Biol 2023;11. Available at: https://pubmed.ncbi.nlm.nih.gov/36819107/ [Accessed September 26, 2023].

26. Anon. Olink Data normalization and standardization.

27. Anon. Facilitate Analysis of Proteomic Data from Olink [R package OlinkAnalyze version 3.5.1]. 2023. Available at: https://CRAN.R-project.org/package=OlinkAnalyze [Accessed October 11, 2023].

28. Storey J, Bass A, Dabney A, Robinson D. qvalue: Q-value estimation for false discovery rate control. 2023.

29. Wu T, Hu E, Xu S, et al. clusterProfiler 4.0: A universal enrichment tool for interpreting omics data. Innovation (Cambridge (Mass)) 2021;2. Available at: https://pubmed.ncbi.nlm.nih.gov/34557778/ [Accessed April 4, 2023].

30. Anon. Chapter 15 Visualization of functional enrichment result | Biomedical Knowledge Mining using GOSemSim and clusterProfiler. Available at: https://yulab-smu.top/biomedical-knowledge-mining-book/enrichplot.html [Accessed April 4, 2023].

31. Wei T, Simko V. R package “corrplot”: Visualization of a Correlation Matrix. 2021.

32. Wickham H. ggplot2: Elegant Graphics for Data Analysis. 2016.

33. Cipriani V, Lorés-Motta L, He F, et al. Increased circulating levels of Factor H-Related Protein 4 are strongly associated with age-related macular degeneration. Nat Commun 2020;11. Available at: https://pubmed.ncbi.nlm.nih.gov/32034129/ [Accessed September 7, 2023].

34. Lorés-Motta L, van Beek AE, Willems E, et al. Common haplotypes at the CFH locus and low-frequency variants in CFHR2 and CFHR5 associate with systemic FHR concentrations and age-related macular degeneration. Am J Hum Genet 2021;108:1367–1384. Available at: https://pubmed.ncbi.nlm.nih.gov/34260947/ [Accessed September 7, 2023].

35. den Hollander AI, Mullins RF, Orozco LD, et al. Systems genomics in age-related macular degeneration. Exp Eye Res 2022;225. Available at: https://pubmed.ncbi.nlm.nih.gov/36108770/ [Accessed September 7, 2023].

36. Kaye R, Chandra S, Sheth J, et al. Central serous chorioretinopathy: An update on risk factors, pathophysiology and imaging modalities. Prog Retin Eye Res 2020;79. Available at: https://pubmed.ncbi.nlm.nih.gov/32407978/ [Accessed September 7, 2023].

37. Schellevis RL, Van Dijk EHC, Breukink MB, et al. Role of the Complement System in Chronic Central Serous Chorioretinopathy: A Genome-Wide Association Study. JAMA Ophthalmol 2018;136:1128–1136. Available at: https://pubmed.ncbi.nlm.nih.gov/30073298/ [Accessed September 7, 2023].

38. de Groot EL, Ossewaarde-van Norel J, de Boer JH, et al. Association of Risk Variants in the CFH Gene With Elevated Levels of Coagulation and Complement Factors in Idiopathic Multifocal Choroiditis. JAMA Ophthalmol 2023;141. Available at: https://pubmed.ncbi.nlm.nih.gov/37410486/ [Accessed September 7, 2023].

39. Ferrara DC, Merriam JE, Freund KB, et al. Analysis of major alleles associated with age-related macular degeneration in patients with multifocal choroiditis: strong association with complement factor H. Arch Ophthalmol 2008;126:1562–1566. Available at: https://pubmed.ncbi.nlm.nih.gov/19001225/ [Accessed September 7, 2023].

40. Ferkingstad E, Sulem P, Atlason BA, et al. Large-scale integration of the plasma proteome with genetics and disease. Nat Genet 2021;53:1712–1721. Available at: https://pubmed.ncbi.nlm.nih.gov/34857953/ [Accessed September 7, 2023].

41. Den Hollander AI, Ten Brink JB, De Kok YJM, et al. Mutations in a human homologue of Drosophila crumbs cause retinitis pigmentosa (RP12). Nat Genet 1999;23:217–221. Available at: https://pubmed.ncbi.nlm.nih.gov/10508521/ [Accessed September 7, 2023].

42. Kousal B, Dudakova L, Gaillyova R, et al. Phenotypic features of CRB1-associated early-onset severe retinal dystrophy and the different molecular approaches to identifying the disease-causing variants. Graefes Arch Clin Exp Ophthalmol 2016;254:1833–1839. Available at: https://pubmed.ncbi.nlm.nih.gov/27113771/ [Accessed September 7, 2023].

43. Lopes da Costa B, Kolesnikova M, Levi SR, et al. Clinical and Therapeutic Evaluation of the Ten Most Prevalent CRB1 Mutations. Biomedicines 2023;11. Available at: https://pubmed.ncbi.nlm.nih.gov/36830922/ [Accessed October 4, 2023].

44. Whaley K, Ruddy S. Modulation of C3b hemolytic activity by a plasma protein distinct from C3b inactivator. Science 1976;193:1011–1013. Available at: https://pubmed.ncbi.nlm.nih.gov/948757/ [Accessed September 7, 2023].

45. Eberhardt HU, Buhlmann D, Hortschansky P, et al. Human factor H-related protein 2 (CFHR2) regulates complement activation. PLoS One 2013;8. Available at: https://pubmed.ncbi.nlm.nih.gov/24260121/ [Accessed September 7, 2023].

46. Cipriani V, Tierney A, Griffiths JR, et al. Beyond factor H: The impact of genetic-risk variants for age-related macular degeneration on circulating factor-H-like 1 and factor-H-related protein concentrations. Am J Hum Genet 2021;108:1385–1400. Available at: https://pubmed.ncbi.nlm.nih.gov/34260948/ [Accessed September 7, 2023].

47. Cserhalmi M, Papp A, Brandus B, et al. Regulation of regulators: Role of the complement factor H-related proteins. Semin Immunol 2019;45. Available at: https://pubmed.ncbi.nlm.nih.gov/31757608/ [Accessed September 7, 2023].

48. Silverman SM, Ma W, Wang X, et al. C3- and CR3-dependent microglial clearance protects photoreceptors in retinitis pigmentosa. J Exp Med 2019;216:1925–1943.

49. Wang S, Du L, Yuan S, Peng GH. Complement C3a receptor inactivation attenuates retinal degeneration induced by oxidative damage. Front Neurosci 2022;16. Available at: https://pubmed.ncbi.nlm.nih.gov/36110094/ [Accessed September 7, 2023].

50. Hoh Kam J, Lenassi E, Malik TH, et al. Complement component C3 plays a critical role in protecting the aging retina in a murine model of age-related macular degeneration. Am J Pathol 2013;183:480–492. Available at: https://pubmed.ncbi.nlm.nih.gov/23747511/ [Accessed September 7, 2023].

51. Wykoff CC, Hershberger V, Eichenbaum D, et al. Inhibition of Complement Factor 3 in Geographic Atrophy with NGM621: Phase 1 Dose-Escalation Study Results. Am J Ophthalmol 2022;235:131–142. Available at: https://pubmed.ncbi.nlm.nih.gov/34509438/ [Accessed September 7, 2023].

52. Yu C, Roubeix C, Sennlaub F, Saban DR. Microglia versus Monocytes: Distinct Roles in Degenerative Diseases of the Retina. Trends Neurosci 2020;43:433–449. Available at: https://pubmed.ncbi.nlm.nih.gov/32459994/ [Accessed September 7, 2023].

53. Guo M, Schwartz TD, Dunaief JL, Cui QN. Myeloid cells in retinal and brain degeneration. FEBS J 2022;289:2337–2361. Available at: https://pubmed.ncbi.nlm.nih.gov/34478598/ [Accessed September 7, 2023].

54. Ong SLM, de Vos IJHM, Meroshini M, et al. Microfibril-associated glycoprotein 4 (Mfap4) regulates haematopoiesis in zebrafish. Sci Rep 2020;10. Available at: https://pubmed.ncbi.nlm.nih.gov/32678226/ [Accessed September 7, 2023].

55. Rajabian F, Arrigo A, Bianco L, et al. Optical Coherence Tomography Angiography in CRB1-Associated Retinal Dystrophies. J Clin Med 2023;12. Available at: https://pubmed.ncbi.nlm.nih.gov/36769743/ [Accessed September 7, 2023].

56. Murro V, Mucciolo DP, Sodi A, et al. Retinal capillaritis in a CRB1-associated retinal dystrophy. Ophthalmic Genet 2017;38:555–558. Available at: 10.1080/13816810.2017.1281966.

57. Alonso F, Dong Y, Li L, et al. Fibrillin-1 regulates endothelial sprouting during angiogenesis. Proc Natl Acad Sci U S A 2023;120. Available at: https://pubmed.ncbi.nlm.nih.gov/37252964/ [Accessed September 7, 2023].

58. Talib M, van Schooneveld MJ, van Genderen MM, et al. Genotypic and Phenotypic Characteristics of CRB1-Associated Retinal Dystrophies: A Long-Term Follow-up Study. Ophthalmology 2017;124:884–895. Available at: https://pubmed.ncbi.nlm.nih.gov/28341475/ [Accessed February 21, 2023].

59. Bevitt DJ, Mohamed J, Catterall JB, et al. Expression of ADAMTS metalloproteinases in the retinal pigment epithelium derived cell line ARPE-19: Transcriptional regulation by TNFα. Biochimica et Biophysica Acta - Gene Structure and Expression 2003;1626:83–91. Available at: https://pubmed.ncbi.nlm.nih.gov/12697333/ [Accessed September 7, 2023].

60. Bushehri A, Zare-Abdollahi D, Alavi A, et al. Identification of PROS1 as a Novel Candidate Gene for Juvenile Retinitis Pigmentosa. Int J Mol Cell Med 2019;8:179–190. Available at: https://pubmed.ncbi.nlm.nih.gov/32489947/ [Accessed September 7, 2023].

61. Nguyen XTA, Talib M, van Schooneveld MJ, et al. CRB1-Associated Retinal Dystrophies: A Prospective Natural History Study in Anticipation of Future Clinical Trials. Am J Ophthalmol 2022;234:37–48. Available at: https://pubmed.ncbi.nlm.nih.gov/34320374/ [Accessed October 3, 2023].

62. Boon N, Lu X, Andriessen CA, et al. AAV-mediated gene augmentation therapy of CRB1 patient-derived retinal organoids restores the histological and transcriptional retinal phenotype. Stem Cell Reports 2023;18:1123–1137. Available at: https://pubmed.ncbi.nlm.nih.gov/37084726/ [Accessed September 7, 2023].

63. da Costa BL, Li Y, Levi SR, et al. Generation of CRB1 RP Patient-Derived iPSCs and a CRISPR/Cas9-Mediated Homology-Directed Repair Strategy for the CRB1 c.2480G>T Mutation. Adv Exp Med Biol 2023;1415:571–576. Available at: https://pubmed.ncbi.nlm.nih.gov/37440088/ [Accessed September 7, 2023].

64. Nguyen X-T-A, Moekotte L, Plomp AS, et al. Retinitis Pigmentosa: Current Clinical Management and Emerging Therapies. Int J Mol Sci 2023;24:7481. Available at: https://www.mdpi.com/1422-0067/24/8/7481 [Accessed April 19, 2023].

65. Gange WS, Sisk RA, Besirli CG, et al. Perifoveal Chorioretinal Atrophy after Subretinal Voretigene Neparvovec-rzyl for RPE65-Mediated Leber Congenital Amaurosis. Ophthalmol Retina 2022;6:58–64. Available at: https://pubmed.ncbi.nlm.nih.gov/33838313/ [Accessed September 7, 2023].

66. Reichel FF, Seitz I, Wozar F, et al. Development of retinal atrophy after subretinal gene therapy with voretigene neparvovec. Br J Ophthalmol 2023;107. Available at: https://pubmed.ncbi.nlm.nih.gov/35609955/ [Accessed September 7, 2023].

